# The cost of diarrhoea: a household perspective from seven countries in the Global Enteric Multicenter Study (GEMS)

**DOI:** 10.1101/2025.01.24.25321093

**Authors:** Md Fuad Al Fidah, Md Ridwan Islam, Rukaeya Amin, Sharika Nuzhat, Tahmeed Ahmed, ASG Faruque

## Abstract

**Background:** The burden of diarrhoeal diseases is considerable in South Asia, as well as in sub-Saharan Africa (SSA). Its economic impact is more profound in resource-limited settings like low- and middle-income countries (LMICs). In this study, we aimed to estimate the direct and indirect cost reported by the caregivers of participants from seven LMICs.

**Methods:** The current study used data from the multicenter, Global Enteric Multicenter Study (GEMS) which enrolled under-5 children (U5C). This prospective case-control study was conducted in 7 sites, all of them located in LMICS. Cost data was collected from the caregivers and after adjustment for inflation, were converted into International Dollar (I$). Quantile regression models were developed after adjusting for age, sex and country.

**Results:** This study analyzed data from 4,592 participants. The median (IQR) total direct cost (TDC) and total indirect cost (TIC) were 8.4 (11.0) I$ and 10.2 (14.3) I$, respectively. Statistically significant differences were found across continents for multiple variables. The highest median TDC and TIC was in Bangladesh (13.6 I$ and 23.2 I$ respectively), while Mozambique reported the lowest (0.4 I$ and 4.9 I$ respectively) with medication accounting for 60.9% of TDC. Quantile regression analysis showed TDC was positively associated with factors like family size, urban residence, moderate-to-severe disease, caregiver education, and use of rehydration methods, while treated drinking water and overweight status were negatively associated. TIC was significantly associated with seeking prior care.

**Conclusion:** The indirect cost of diarrhoea was higher than the direct cost which indicates the impact of lost productivity due to the disease. Bolstering the healthcare financing systems, ensuring affordable medication, promoting WASH initiative and timely healthcare-seeking can reduce the economic burden.

## Introduction

Among children under the age of 5 years, diarrhoea is still one of the leading causes of mortality and morbidity.[1] Due to being preventable and treatable with measures like oral rehydration saline (ORS), the overall mortality due to diarrhoea has seen significant decline (approximately 55%) since the 2000’s.[2] The burden of diarrhoeal diseases is considerable in South Asia, as well as in sub-Saharan Africa (SSA). The countries from these to geographic locations are responsible for 90% of the global death due to diarrhoea.[3]

Apart from mortality, the economic impact of the disease on the households and the health system is also crucial. In resource-limited settings like LMICs, the impact is more profound.[2] Studies have proven that this economic burden even negatively impacts the household’s ability to access diarrhoeal treatment.[2,4] Moreover, diarrhoea leads to malnutrition which may result in death. [2] The disease also causes financial strains on the families as loss of productivity is expected for the caregiver of the sick child.[5]

Several studies have aimed to estimate the household cost and the economic implication of the diarrhoea among under-5 children in various countries.[5] A study conducted in Bangladesh reported that on average, households spend 26.2 International dollars (I$) in direct cost for treating diarrhoea before reporting to a diarrhoeal diseases hospital.[5] As reported by Das et al., the total cost of invasive enteritis among under-5 children in Bangladesh was almost similar based on pathogens namely Shigella (4.17 USD) and Campylobacter (3.49 USD).[6] Another report based on the data from the Global Enteric Multicenter Study (GEMS) study also estimated the household costs of diarrhoea by etiology in seven countries revealed that the household out-of-pocket (OOP) costs were higher in Mali, whereas differences due to etiology within countries were not statistically significant.[7] However, comprehensive data related to direct and indirect cost due to diarrhoea in South Asia and SSA is scare.

Cost-of-illness (COI) can act a useful guideline for policy makers in prioritizing, selecting and scaling up of interventions.[2] The GEMS study was conducted on 7 sites where 3 countries from South Asia and 4 from SSA were included.[8] Between 2007 and 2011, under-5 children (U5C) with diarrhoeal disease were enrolled in the study. During enrollment, information were collected from the parents or caregivers.[7] In this study, we aimed to estimate the direct and indirect cost reported by the caregivers of participants from the GEMS study.

## Methods

### Data overview

We used data from the Global Enteric Multicenter Study (GEMS). The details of the GEMS can be found elsewhere.[8–10] The data was collected from ClinEpiDB, an open-access online resource for clinical and epidemiologic studies, after signing up and receiving necessary permission.[11] Briefly, the GEMS enrolled children under the age of 5 years in the study from 7 sites. In South Asia 3 countries were selected (Bangladesh, India and Pakistan); additionally, 4 countries from Sub-Saharan Africa were also selected as study sites (Kenya, Mali, Mozambique, and the Gambia) (Fig 1). This prospective case-control study was conducted from December 2007 to March 2011. Children aged <5 years residing within each of the sites demographic surveillance systems (DSS) were eligible for the study. Each site had sentinel health centers where children were assessed and matched based on the study’s inclusion criteria.

**Fig 1:**
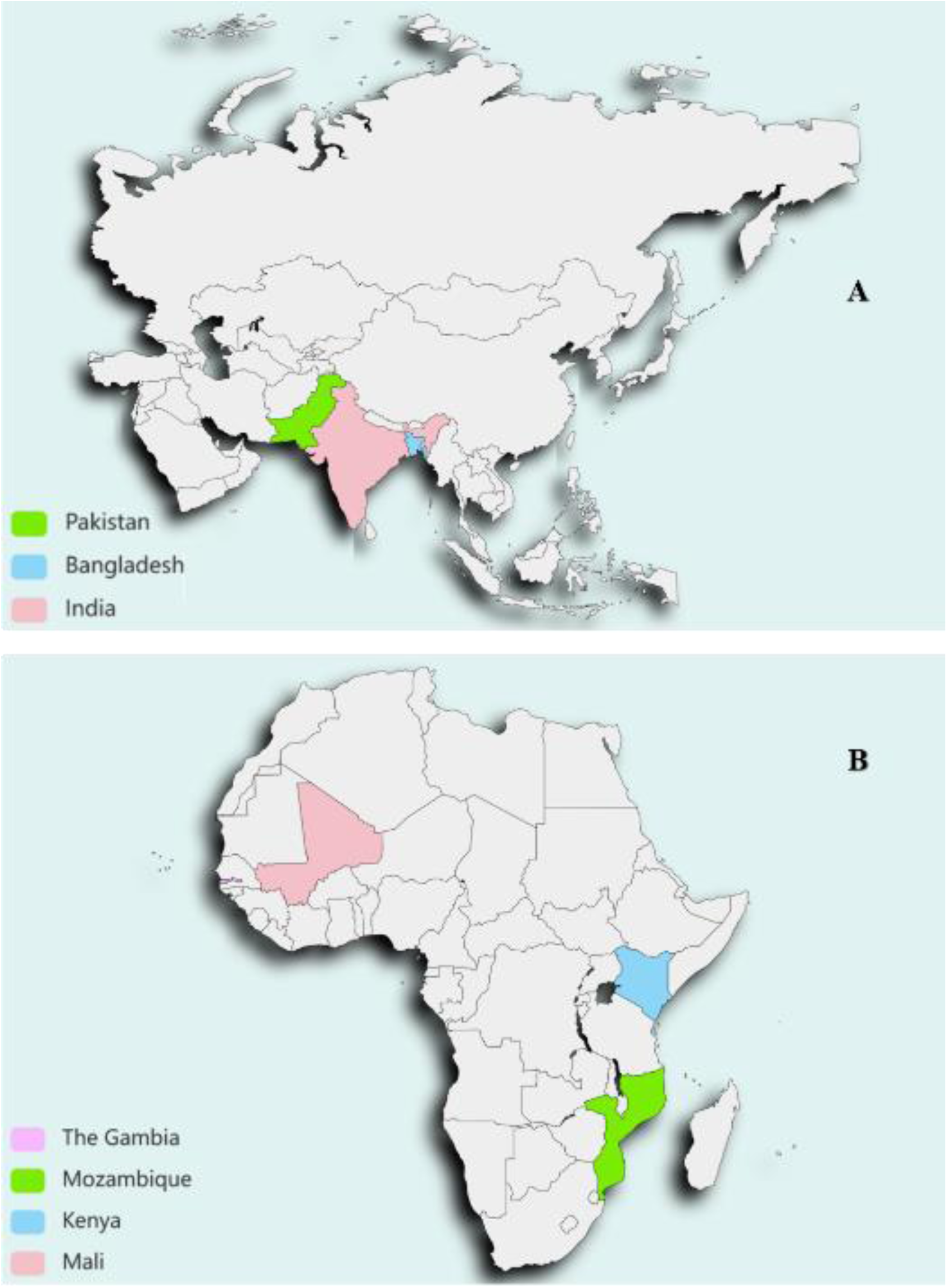
Study sites for the GEMS. A) 3 study sites in South Asia; B) 4 study sites in Sub-Saharan Africa

The GEMS enrolled 14242 participants in total with 5844 of them being cases and 8398 were matched controls. The ‘cases’ were children aged <60 months who reported to the sentinel health center with acute moderate-to-severe diarrhoeal disease (MSD) and controls were children without MSD. We considered 4592 children aged <60 months for our analysis. Only cases with non-zero total costs were considered following literature review.[6] The procedure used to select children for this investigation is described in detail in Fig 2.

**Fig 2:**
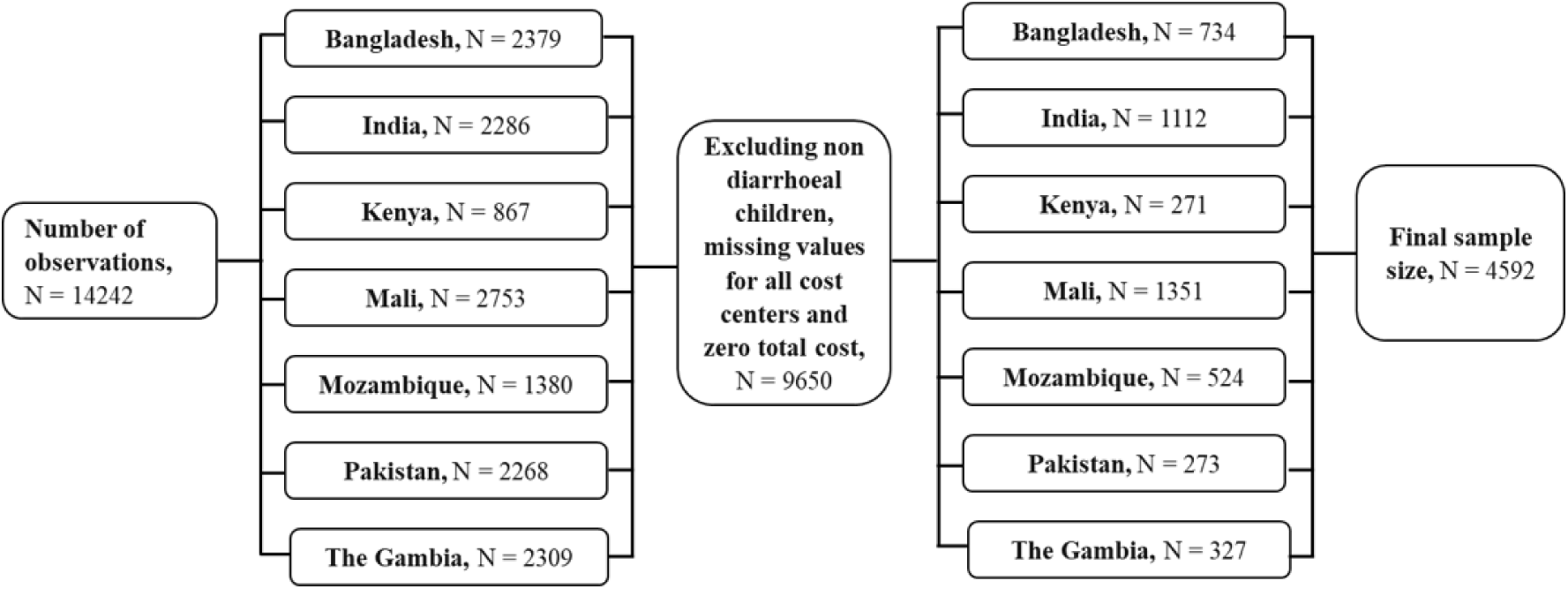
Selection of children <60 months for analysis

### Operational definitions and measurements

#### Outcome variables

We used the healthcare consumer’s perspective while calculating costs. The outcome variables of our study were total direct cost (TDC) and total indirect cost (TIC). All costs were calculated in the participant’s local currency, then adjusted for inflation and converted into International dollars (I$) for better comparison.

#### Total direct cost (TDC)

The direct cost is defined as the amount of money spend for medical management of the disease and it includes the cost of drugs, consultation, diagnostic, patient transportation, hospital cost, food cost etc.[12] Studies have suggested that there are more the 30 cost centers that can be considered for calculating direct cost. However, some of these cost centers may not be appropriate for every patient.[5,12] We inquired about the expenses incurred for prior healthcare seeking as well as the current healthcare seeking in local currency. The amount of expenses for current care seeking was asked after discharge from the hospital which were self-reported by the caregivers. In our study, total direct cost was calculated by summing the cost for direct medical cost for previously sought care for diarrhoea (pharmacy, consultation irrespective of type of provider, medication cost, hospital cost, other medical expenses), direct non-medical cost for previously sought care for diarrhoea (transport cost for pharmacy, consultation irrespective of type of provider, hospital visit, buying drugs, other transportation expenses), direct medical cost for current care (consultation cost, hospitalization cost, medication cost, diagnostic cost, other medical cost) and direct non-medical cost for current care (food expenses, hospital expenses for card, soap etc., transportation cost for the patient and other household members).

#### Total indirect cost (TIC)

Indirect costs are expenses that arise from the consequences of an illness, rather than from direct medical treatment. These can include lost income, decreased productivity, and additional expenses like home care or childcare.[12] We calculated indirect costs using days of absenteeism from work for the care givers. Salary loss due to caregiving illness was calculated using the average daily income of the caregiver in local currency.[6,13,14] For the loss of half a morning or afternoon, 0.25 days were considered. Similarly, for a morning or afternoon it was 0.50 days, a morning and afternoon it was 1.00 days and for less than half a morning or afternoon it was 0.00 days. The loss of time was also calculated in the same fashion if any other caregiver took care of the child.

#### Independent variables

The independent variables we considered for this study include age (in months), age group, number of family members, number of U5C, sex (male/female), residence (urban/rural), died (yes/no), MSD (yes/no), previously sought care (yes/no), care by licensed practitioner (yes/no), primary caregiver (mother/others), level of education of primary caregiver (informal or up to primary/ primary completed/ secondary completed/ post-secondary), rehydration method used (none/oral/IV), wealth index (poorest/poor/middle/rich/richest), treated drinking water (yes/no), stunting (yes/no), wasting (yes/no), underweight (yes/no), overweight (yes/no) and components of TDC (pharmacy cost/ consultation cost/ medication cost/ healthcare center cost/ diagnostic cost/ other cost).

#### Residence

We considered the location of the study site to determine whether it is urban or rural. As reported by Levine et al., Mali, India and Pakistan were considered as urban setting and the four other countries as rural setting.[15]

#### MSD

We defined MSD as diarrhoea with the presence of at least 1 of these symptoms sunken eyes, loss of skin turgor, intravenous hydration administered or prescribed, dysentery, admission or advised admission to hospital. [15]

#### Wealth index

It is a method that is used by various survey studies like Multiple Indicator Cluster Survey or Demographic and health surveys. Using this approach, the participants can be classified into 5 quantiles on the premise that the possession of households assets and amenities indicates the households relative economic position. [16,17]

#### Stunting

U5C with height/length-for-age z score of <-2 standard deviations (SD) were considered as stunted. [18]

#### Wasting

U5C with weight-for-height z score of <-2 standard deviations (SD) were considered as wasted. [18]

#### Underweight

U5C with weight-for-age z score of <-2 standard deviations (SD) were considered as underweight. [18]

#### Overweight

U5C with BMI-for-age z score of >1 standard deviations (SD) were considered as overweight. [18]

#### Adjusting for inflation

Inflation in a phenomenon where the same amount money loses its purchasing ability over the years, mostly due to an increase in price. The Consumer Price Index (CPI) is used to tracks the impact of inflation. [19] The following formula is used to adjust expenses or cost for inflation [20]:

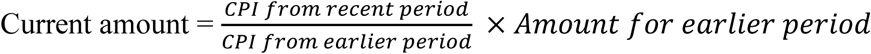

We considered 2022 the recent period for all cost. The ‘general’ CPI for each country was used to adjust for inflation. The CPIs of each center are shown in S1 Table.[21]

#### Conversion of currency

For economic studies, costs are adjusted for inflation and reported in US dollars or international dollars (I$).[5] The I$ is a hypothetical unit of currency designed to account for differences in relative prices across various contexts. For example, I$1 would purchase a comparable amount of goods and services in the country of interest as 1 USD would in the United States.[5] The current study used the following formula for conversion of the local currency, I$[19]:

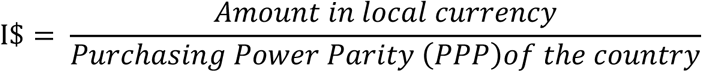

The PPP in 2022 for the study sites are presented in S1 Table.[22]

### Statistical analysis

Data analysis was done based on the type of variable. Normality of continuous data was checked using Shapiro-Franscia test and were presented using median (IQR). The categorical data was presented using frequency and percentage. We used Mann-Whitney U test, Kruskal-Wallis test (Dunn test as post-hoc estimation) and chi-square test to investigate association between variables where applicable. Quantile regression analysis was conducted to identify predictor variables for TDC and TIC after adjusting for age, sex and country. Only adjusted coefficients for significant predictors were reported with their corresponding 95% CI (confidence interval). All statistical tests were two-sided, with a significance level of α=0.05 and were conducted using STATA 17 (StataCorp LLC, Texus, USA).

### Ethical consideration

The study analysed data from the GEMS study which is available on registration and approval from the website: https://clinepidb.org/ce/app. Prior to the implementation of the Global Enteric Multicenter Study, the case management protocols, consent documentation, case report forms, field procedures, and other research-supporting materials underwent formal authorization by the Institutional Review Board (IRB) of the University of Maryland School of Medicine in Baltimore, MD. Additionally, IRB approval was secured by the committees and collaborating partners overseeing operations at the seven participating institutions. These institutions included the International Centre for Diarrhoeal Disease Research, Bangladesh (icddr,b) in Bangladesh; the National Institute of Cholera and Enteric Diseases in India; Aga Khan University in Pakistan; the Medical Research Council Unit in The Gambia; the CDC/Kenya Medical Research Institute Research Station in Kenya; the Centre pour le Développement des Vaccins du Mali in Mali; and the Centro de Investigação em Saúde de Manhiça in Mozambique. Informed consent forms, signed by the parents or guardians of participating children (both cases and healthy controls), were obtained prior to enrollment.

## Results

The following study analyzed data from 4592 participants. The process of selecting the participants is presented in Fig 2. The total direct cost (TDC) was reported by 4592 participants and 430 participants reported total indirect cost (TIC). The median (IQR) TDC and TIC were 8.4 (11.0) and 10.2 (14.3) I$ respectively. There was a significant (p-value=0.015) positive correlation (Spearman’s rho=0.012) between TDC and TIC (Fig 3).

**Fig 3:**
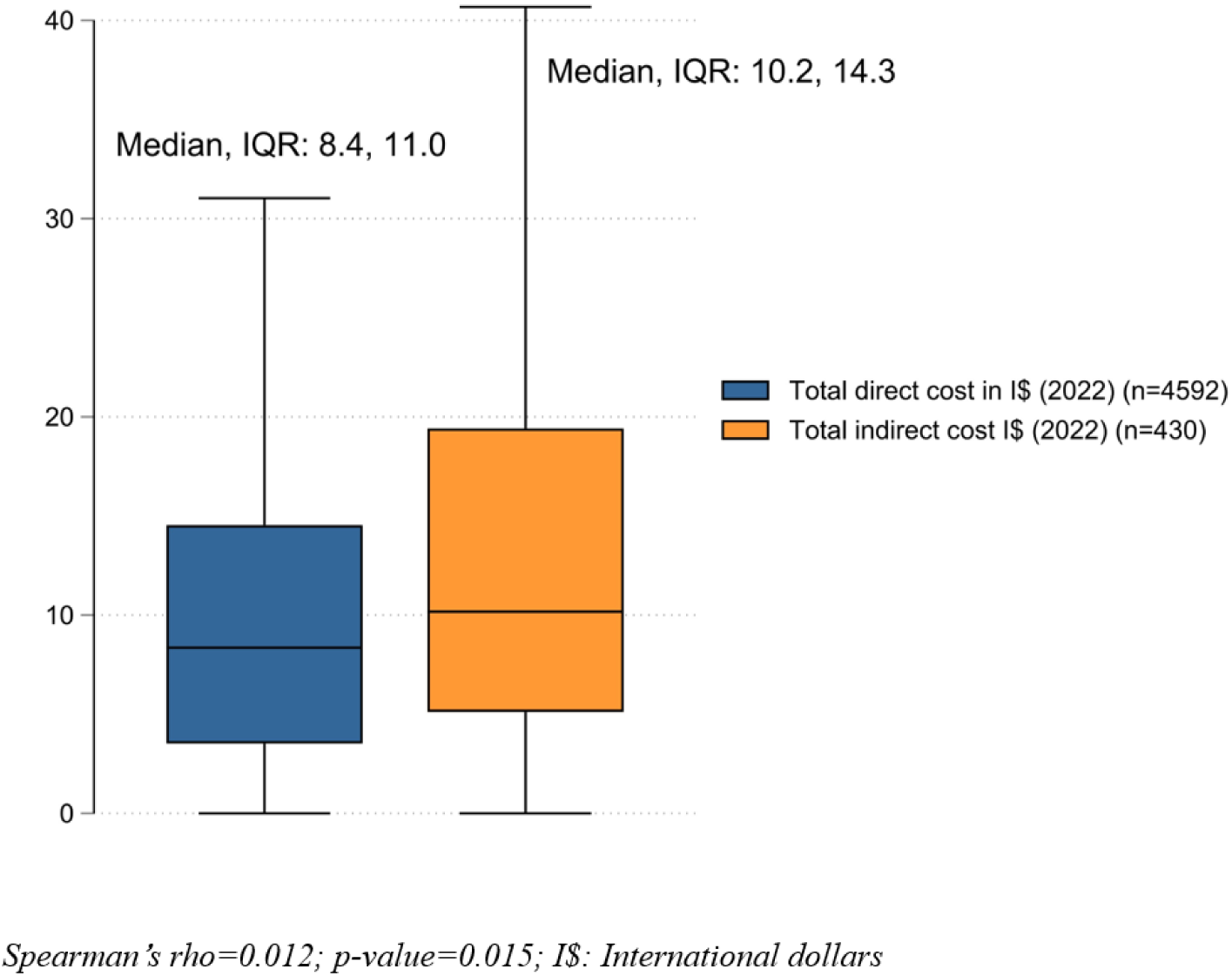
Median total direct cost and total indirect cost in I$

The distribution of independent variables by continent is presented in Table 1. The median (IQR) age of the participants was 14 (16) months. Most of them were male (54.6%) and resided in urban areas (59.6%). Statistically significant association was found between continent and no. of family numbers (p-value<0.001), no. of children under 5 years (p-value<0.001), residence (p-value<0.001), death (p-value=0.007), previously sought care (p-value<0.001), care by licensed practitioner (p-value<0.001), primary caregiver (p-value<0.001), level of education of the primary caregiver (p-value<0.001), rehydration method used (p-value<0.001), wealth index (p-value<0.001), treated drinking water (p-value<0.001), stunting (p-value<0.001), underweight (p-value<0.001), overweight (p-value<0.001) and TDC (p-value<0.001) (Table 1).

**Table 1:** Distribution of independent variables by continent (n=4592)

| Variables | Continent |  |  |  |
| --- | --- | --- | --- | --- |
|  | Total<br>n (%) | South Asia<br>n (=2119)<br>(%= 46.2) | Sub-Saharan<br>Africa<br>n (=2473) (%=53.8) | p-value |
| Age (in months)* | 14 (16) | 14 (16) | 14 (17) | 0.268 |
| Age group (in months) |  |  |  | 0.223 |
| 0-11 | 1822 (39.7) | 689 (41.0) | 953 (38.5) |  |
| 12-23 | 1524 (33.2) | 684 (32.3) | 840 (34.0) |  |
| 24-59 | 1246 (27.1) | 566 (26.7) | (27.5) |  |
| No. of family members* | 7 (7) | 6 (4) | 10 (16) | <0.001 |
| No. U5C* | 2 (2) | 1 (1) | 2 (3) | <0.001 |
| Sex (male), n (%) | 2507 (54.6) | 1179 (55.6) | 1328 (53.7) | 0.188 |
| Residence (urban), n (%) | 2736 (59.6) | 1385 (65.4) | 1351 (54.6) | <0.001 |
| Died (yes), n (=4351) (%) | 31 (0.7) | 7 (0.4) | 24 (1.0) | 0.007 |
| MSD (yes), n (%) | 2214 (48.2) | 996 (47.0) | 1217 (49.2) | 0.135 |
| Previously sought care (yes),<br>n (%) | 1355 (29.5) | 908 (42.9) | 407 (18.1) | <0.001 |
| Care by licensed practitioner<br>(yes) , n (=1355) (%) | 175 (12.9) | 168 (18.5) | 7 (1.6) | <0.001 |
| Primary caregiver (mother), n<br>(%) | 4402 (95.9) | 2087 (98.5) | 2315 (93.6) | <0.001 |
| Level of education of primary caregiver (n=4588) |  |  |  |  |
| Informal or up to primary | 2572 (56.1) | 720 (34.0) | 1852 (75.0) | <0.001 |
| Primary completed | 1562 (34.1) | 1088 (51.3) | 474 (19.2) |  |
| Secondary completed | 343 (7.5) | 224 (10.6) | 119 (4.8) |  |
| Post-secondary | 111 (2.4) | 87 (4.1) | 24 (1.0) |  |
| Rehydration method used |  |  |  |  |
| None | 753 (16.4) | 91 (4.3) | 662 (26.8) | <0.001 |
| Oral | 3625 (78.9) | 1953 (92.2) | 1672 (67.6) |  |
| IV | 214 (4.7) | 75 (3.5) | 139 (5.6) |  |
| Wealth index (n=4591) |  |  |  |  |
| Poorest | 981 (21.4) | 451 (21.3) | 530 (21.4) | <0.001 |
| Poor | 961 (20.9) | 490 (23.1) | 471 (19.1) |  |
| Middle | 856 (18.6) | 349 (16.5) | 507 (20.5) |  |
| Rich | 925 (20.2) | 444 (20.9) | 481 (19.5) |  |
| Richest | 868 (18.9) | 385 (18.2) | 483 (19.5) |  |
| **Treated drinking water** (yes),<br>n (%) | 929 (20.2) | 602 (28.4) | 327 (13.2) | <0.001 |
| **Stunting** (yes), n (%) | 1068 (23.3) | 572 (27.0) | 496 (20.1) | <0.001 |
| **Wasting** (yes), n (%) | 672 (14.6) | 303 (14.3) | 369 (14.9) | 0.552 |
| **Underweight** (yes), n (%) | 119 (24.4) | 608 (28.7) | 511 (20.7) | <0.001 |
| **Overweight** (yes), n (%) | 389 (8.5) | 145 (6.8) | 244 (9.9) | <0.001 |
| **Total direct cost (in I\$)\*** | 8.4 (11.0) | 10.3 (7.6) | 5.1 (12.9) | <0.001 |
| **Total indirect cost (in I\$)**<br>(n=430)* | 10.2 (14.3) | 11.3 (13.4) | 10.2 (15.3) | 0.328 |

\*Median (IQR), p-value from Mann-Whitney U test reported, MSD: Moderate-to-severe disease;
IV: Intra-venous; I\$: International dollars

The distribution of the TDC in I$ is presented in Fig 4. The highest median (IQR) TDC was found in Bangladesh (13.6, 9.8 I$) and the lowest was in Mozambique (0.4, 1.2 I$). Significant association was found between country and the TDC (p-value<0.001). Significant difference was also found between the TDC reported by the countries.

**Fig 4:**
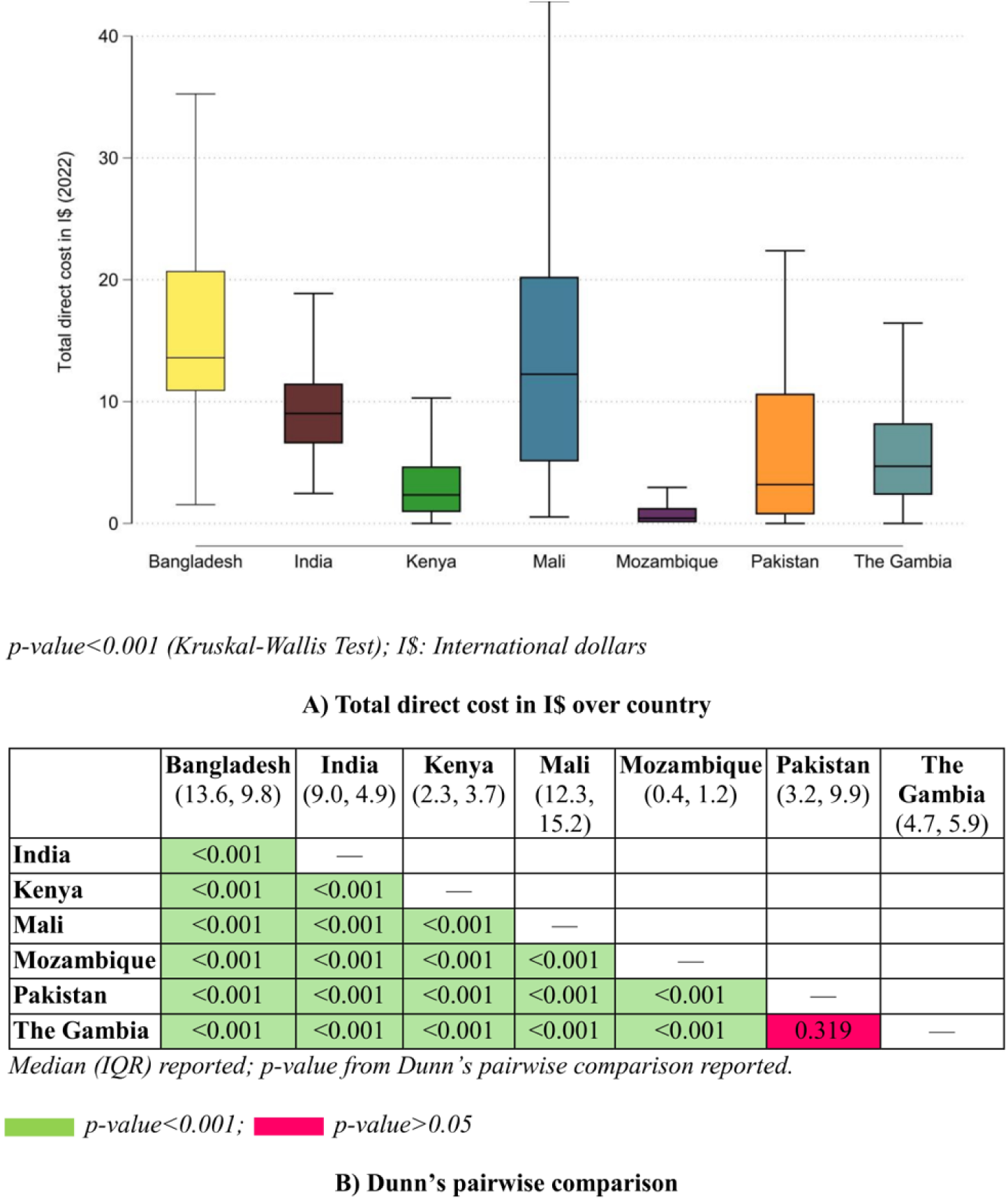
Total direct cost (in I$) across countries

The highest proportion of TDC was due to medication (60.9%), and diagnostic cost was responsible for only 0.7% of the TDC (Supplementary Fig 1).

The maximum median (IQR) TIC was reported by Bangladesh (23.2, 14.5 I$). The Gambia also reported a high median (IQR) TIC of 19.7 (22.9) I$. However, the minimum was reported by Mozambique (7.0, 4.9 I$). The TIC was significantly associated with the country (p-value<0.001) (Fig 5).

**Fig 5:**
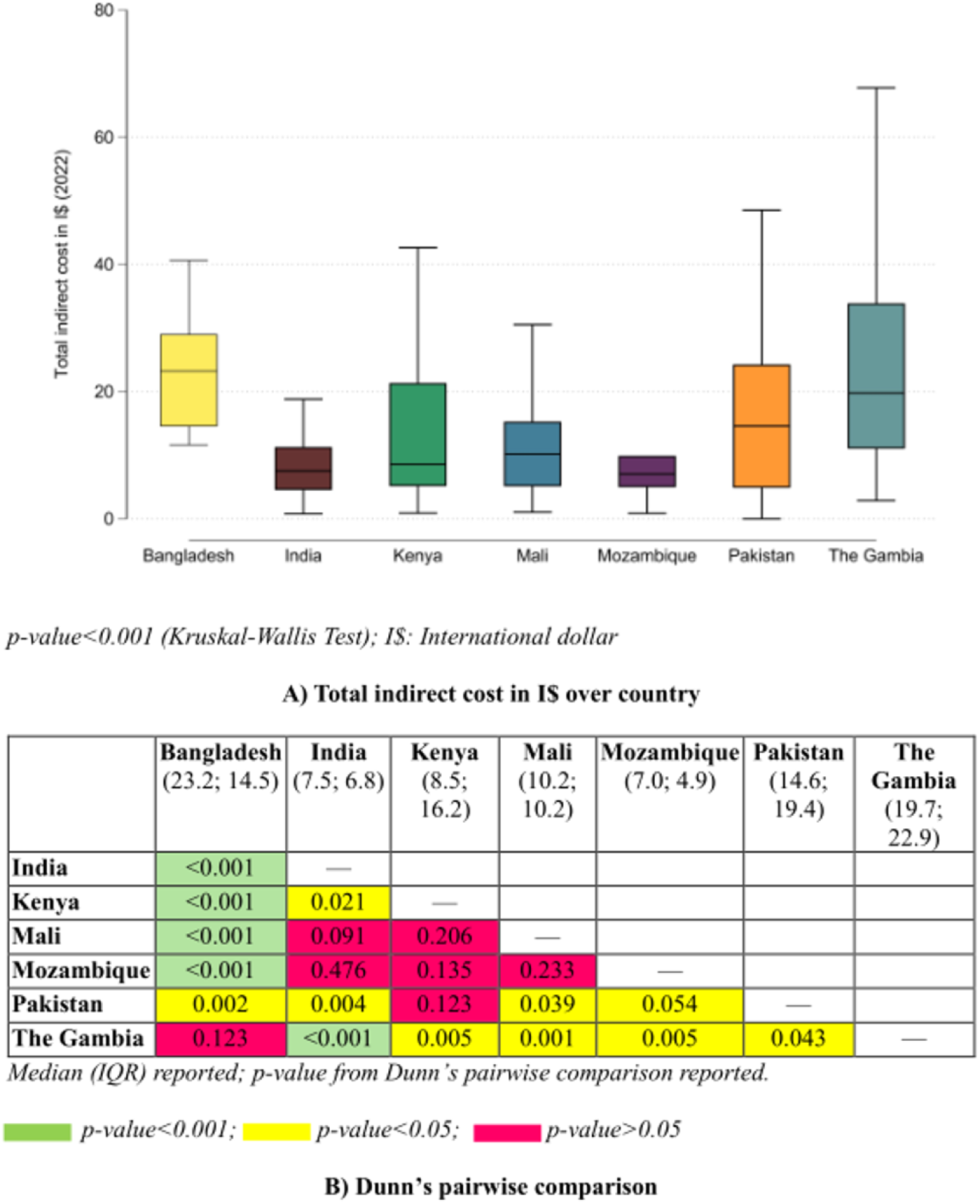
Total indirect cost (in I$) across countries (n=430)

Bivariate quantile regression models were developed to identify independent variables associated with TDC. Models were adjusted for age in months, sex and country. We found significant positive association between TDC and no. of family members, no. of U5C, urban residence, having MSD, previously seeking care, post-secondary level of education of the primary caregiver, using oral or IV rehydration method, being wasted and being underweight. However, primary completed level of education of the primary caregiver, treatment of drinking water, loose or water stool and having overweight had significant negative association (Table 2).

**Table 2:** Quantile regression analysis for association between total direct cost and predictor variables.

| Variable | Unadjusted coef. |  | Adjusted coef. |  |
| --- | --- | --- | --- | --- |
|  | Coef. (95% CI: lower; upper) | p-value | Coef. (95% CI: lower; upper) | p-value |
| **Age (in months)** | -0.01 (-0.03; 0.02) | 0.506 | — | — |
| **Age group (in months)** | Ref: 0-11 |  |  |  |
| 12-23 | 0.5 (-0.2; 1.2) | 0.181 | — | — |
| 24-59 | -0.3 (-1.1; 0.5) | 0.419 | — | — |
| **No. of family members** | 0.00 (-0.02; 0.03) | 0.735 | 0.19 (0.17; 0.21) | <0.001 |
| **No. of U5C** | -0.13 (-0.29; 0.02) | 0.091 | 0.78 (0.67; 0.90) | <0.001 |
| **Sex** | Ref: Female |  |  |  |
| Male | 0.41 (-0.23; 1.05) | 0.212 | — | — |
| **Residence** | Ref: Rural |  |  |  |
| Urban | 3.45 (2.84; 4.07) | <0.001 | 1.56 (1.07; 2.04) | <0.001 |
| **Died (n=4351)** | Ref: No |  |  |  |
| Yes | -3.22 (-6.88; 0.44) | 0.085 | — | — |
| **MSD** | Ref: No |  |  |  |
| Yes | 4.57 (4.13; 4.99) | <0.001 | 3.65 (3.21; 4.09) | <0.001 |
| **Previously sought care** | Ref: No |  |  |  |
| Yes | 8.21 (7.68; 8.73) | <0.001 | 7.18 (6.63; 7.73) | <0.001 |
| **Care by licensed practitioner (n=1355)** | Ref: No |  |  |  |
| Yes | -2.59 (-4.72; -0.45) | 0.018 | — | — |
| **Primary caregiver** | Ref: No |  |  |  |
| Mother | 0.46 (-1.09; 2.01) | 0.561 | — | — |
| **Level of education of primary caregiver (n=4588)** | Ref: Informal or up to primary |  |  |  |
| Primary completed | 2.87 (2.25; 3.49) | <0.001 | -1.12 (-1.66; -0.58) | <0.001 |
| Secondary completed | 2.87 (1.76; 3.98) | <0.001 | — | — |
| Post-secondary | 5.08 (3.21; 6.96) | <0.001 | 1.47 (-0.00; 2.94) | 0.050 |
| **Rehydration method used** | Ref: None |  |  |  |
| Oral | 8.97 (8.25; 9.68) | <0.001 | 6.41 (5.53; 7.29) | <0.001 |
| IV | 10.81 (9.44; 12.19) | <0.001 | 10.33 (8.92; 11.74) | <0.001 |
| **Wealth index (n=4591)** | Ref: Poorest |  |  |  |
| Poor | 0.00 (1.102; 1.02) | >0.999 | — | — |
| Middle | 0.08 (-0.97; 1.12) | 0.884 | — | — |
| Rich | 0.16 (-0.86; 1.19) | 0.754 | — | — |
| Richest | 0.98 (-0.06; 2.03) | 0.064 | — | — |
| **Treated drinking water** | Ref: No |  |  |  |
| Yes | -1.81 (-2.67; -0.94) | <0.001 | -1.46 (-2.00; -0.92) | <0.001 |
| **Stunting** |  | Ref: No |  |  |
| Yes | -0.25 (-1.03; 0.51) | 0.524 | — | — |
| **Wasting** |  | Ref: No |  |  |
| Yes | 2.36 (1.48; 3.25) | <0.001 | 2.57 (1.93; 3.20) | <0.001 |
| **Underweight** |  | Ref: No |  |  |
| Yes | 1.23 (0.49; 1.97) | 0.001 | 1.31 (0.79; 1.83) | <0.001 |
| **Overweight** |  | Ref: No |  |  |
| Yes | -3.29 (-4.42; -2.16) | <0.001 | -2.26 (-3.07; -1.45) | <0.001 |

Variables are adjusted for age, sex and country; MSD: Moderate-to-severe disease; IV: Intra-venous

Adjusted bivariate quantile regression model (adjusted for age in months, sex and country) was developed to examine the association between independent variables and TIC. After adjusting, we found positive significant association between TIC and previously seeking care. However, having loose or watery stool had a significant negative association (Table 3).

**Table 3:** Quantile regression analysis for association between total indirect cost and predictor variables (n=430)

| Variable | Unadjusted coef. |  | Adjusted coef. |  |
| --- | --- | --- | --- | --- |
|  | Coef. (95% CI: lower; upper) | p-value | Coef. (95% CI: lower; upper) | p-value |
| **Age (in months)** | -0.05 (-0.14; 0.03) | 0.223 | — | — |
| **Age group (in months)** |  | Ref: 0-11 |  |  |
| 12-23 | -0.49 (-2.82; 1.85) | 0.682 | — | — |
| 24-59 | -1.23 (-4.04; 1.50) | 0.367 | — | — |
| **No. of family members** | 0.00 (-0.10; 0.10) | >0.999 | — | — |
| **No. U5C** | 0.05 (-0.43; 0.52) | 0.848 | — | — |
| **Sex** |  | Ref: Female |  |  |
| Male | -1.56 (-3.59; 0.47) | 0.133 | — | — |
| **Residence** |  | Ref: Rural |  |  |
| Urban | -0.56 (-2.93; 1.82) | 0.645 | — | — |
| **Died (n=395)** |  | Ref: No |  |  |
| Yes | 5.08 (-1.02; 11.19) | 0.103 | — | — |
| **MSD** |  | Ref: No |  |  |
| Yes | 0.00 (-2.13; 2.14) | >0.999 | — | — |
| **Previously sought care** |  | Ref: No |  |  |
| Yes | 4.26 (2.56; 5.96) | <0.001 | 3.65 (1.57; 5.72) | <0.001 |
| **Care by licensed practitioner** |  | Ref: No |  |  |
| Yes | 2.30 (-3.12; 7.73) | 0.403 | — | — |
| **Primary caregiver** |  | Ref: No |  |  |
| Mother | 0.00 (-4.45; 4.45) | >0.999 | — | — |
| **Level of education of primary caregiver** |  | Ref: Informal or up to primary |  |  |
| Primary completed | 0.00 (-2.05; 2.05) | >0.999 | — | — |
| Secondary completed | 1.09 (-3.03; 5.21) | 0.603 | — | — |
| Post-secondary | 4.84 (-0.96; 10.65) | 0.102 | — | — |
| **Rehydration method used** |  | Ref: None |  |  |
| Oral | 0.28 (-3.20; 3.76) | 0.872 | — | — |
| IV | 0.77 (-3.56; 5.11) | 0.727 | — | — |
| **Wealth index** |  | Ref: Poorest |  |  |
| Poor | 1.44 (-1.89; 4.76) | 0.397 | — | — |
| Middle | 1.44 (-1.39; 4.26) | 0.318 | — | — |
| Rich | 2.53 (-0.40; 5.45) | 0.090 | — | — |
| Richest | 1.44 (-1.65; 4.52) | 0.361 | — | — |
| **Treated drinking water** |  | Ref: No |  |  |
| Yes | 0.00 (-2.16; 2.16) | >0.999 | — | — |
| **Stunting** |  | Ref: No |  |  |
| Yes | 0.00 (-2.42; 2.42) | >0.999 | — | — |
| **Wasting** |  | Ref: No |  |  |
| Yes | 1.12 (-1.60; 3.85) | 0.419 | — | — |
| **Underweight** |  | Ref: No |  |  |
| Yes | 1.09 (-1.11; 3.29) | 0.330 | — | — |
| **Overweight** |  | Ref: No |  |  |
| Yes | 2.54 (-1.00; 6.09) | 0.160 | — | — |

\*Adjusted for age, sex and country; MSD: Moderate-to-severe disease; IV: Intra-venous

## Discussion

In the past few decades, the world has seen a significant decline in the prevalence of pediatric diarrhoeal diseases. However, it still remains one of the major public health concerns, especially in LMICs.[23] It has been reported that children are more susceptible to the disease with an average incidence of 3.2 to 12 episodes per year per child, although most of these episodes are self-limiting.[23] Apart from the obvious physical toll it exerts on the child, the economic strain of diarrhoeal disease on the household is by no means negligible.[2] In this study, we analyzed the data from the GEMS study. It was a multicentered study considering U5C as the participants. The study collected household cost related data from the caregivers. We found that the median total indirect cost was higher than the total direct cost (10.2 I$ vs 8.4 I$). This finding signifies the often-neglected effect of diarrhoeal diseases, i.e., the economic strain it puts on the family due to loss of productivity. Koopmanschap and Rutten have indicated that if any healthcare program produce health benefits quickly, then the impact of that particular program is more impactful.[24] Unfortunately, programs that aims to reduce diarrhoeal burden, often fails to consider this point. Our study found some significant differences regarding the independent variables between the two continents (South Asia and Sub-Saharan Africa). This indicates that although the economic situation between the countries of these two continents are often similar, there are marked differences in the culture and social dynamics.[25] The TDC was significantly higher in Sub-Saharan Africa (12.9 I$) than South Asia (7.6 I$). TIC was also higher in Sub-Saharan Africa, although the difference was not statistically significant. In their study, Asante et al., presents various findings that can account for this fact.[26] Although the region bears a disproportionate share of global disease burden, the allocation of resources to healthcare is quite low. Additionally, in 2016, the per capita health spending in Africa was more than 50 times lower compared to the Organisation for Economic Co-operation and Development countries.[26] Furthermore, the financing systems for healthcare in SSA is largely dependent on high OOP payment with very low government spending.[26]

However, when compared between countries, Bangladesh reported the highest TDC and TIC. When we consider the Per capita total expenditure on health in I$, this finding becomes understandable. During 2011-12, i.e., the period of the GEMS study, Bangladesh spend 66.8 and 67.8 I$ respectively which was the second lowest among all 7 participating countries.[27] It has been reported that OOP payments by the households are responsible for nearly 67% of the total health expenditure.[28] Several reasons like poorly designed health financing system, lack of proper health insurance measures, hidden charges, understaffing etc. are often cited as the root cause of this high OOP rate.[28]

Our study found that the highest proportion of TDC was due to medication while diagnostic cost was the minimum. In developed countries, the government usually provides subsidy for pharmaceuticals. As the cost of medicines are subsidized, the direct cost is not much impacted. On the other hand, in LMICs like Pakistan, the prices of drugs are usually controlled at the level of retail pharmacy.[29] As such, the higher proportion of medication is understandable.

We found that no. of family members and no. of U5C increased the median TDC. The effect of number of family members and other U5C is most likely to be indirect. Several studies have reported family size to be a significant influencer of diarrhoea among U5C.[30,31] One possible reason is the low level of care by the parents and increased risk of transmission.[30]

Residence was an important predictor of TDC where urban households are more likely to have higher cost. The living cost in urban areas is significantly higher when compared to rural areas. Additionally, the cost of care at tertiary level hospitals, which are more prevalent in the urban areas, are significantly higher.[32]

Diarrhoeal diseases can be categorized as less severe disease and moderate to severe disease (MSD) based on level of dehydration, presence of blood in stool and hospital admission.[15] U5C with MSD are usually provided in-patient care. Thobari et al. conducted Health facilities and community survey to estimate the economic burden of U5C in Indonesia where they found that DTC for in-patient cost was significantly higher than out-patient care. Our study reports similar findings. It has been reported that patients with in-patient care requires higher professional fees, diagnostic and medication costs. Additionally, the cost become higher due to the presence of several specialty and subspecialty medical professionals.[32] Another study conducted in Bangladesh reported similar findings were MSD was associated with direct medical and direct non-medical cost.[5] As ORS or IV is used as the rehydration method for MSD, the positive association between rehydration method and TDC found in our study is also understandable.

We found that, children with mothers who completed primary education reported lower cost. However, when the maternal educational level is post-secondary, the cost increases. In LMICs, mothers are usually considered as the primary caregivers. Education is known to have significant impact on the maternal care-seeking behavior for their child’s illness.[5] Reports suggest that when the mother is highly educated, their tendency to more frequent care-seeking also increases, leading to higher expenditure.[5]

Diarrhoea is a water-borne disease, which is reported to be linked with the quality of the drinking water.[23,33] In a cluster randomized controlled trial, Solomon et al. reported that treating drinking water considerably decreases the diarrhoeal episodes.[34] According the WHO, 94.0% of diarrhoeal incidence can be prevented simply by modifying the environment, improving hygiene and sanitation, and making clean water more available.[34] We also found that consuming treated drinking water negatively impacted TDC.

When the child’s condition does not improve, mothers often seek additional consultations. In LMICs, the population often seek care from informal caregivers rather than professional doctors.[35] However, this causes multiple negative impact on patient. These informal caregivers doesn’t have formal care or education which exacerbates the disease leading to increased direct and indirect cost. Moreover, seeking care from multiple sources puts additional financial strain. Our findings also indicate that seeking previous care has caused increased TDC and TIC.

It has been suggested by multiple studies that malnutrition can increased the frequency and duration of diarrhoeal illness.[36,37] Specially wasting is reported to be a strong predictor for prolonged illness.[37] Several mechanisms are suggested for explaining this phenomenon like poor appetite, malabsorption of nutrients, hastening of intestinal transit time etc. Our study has also reported a positive association between wasting, underweight and TDC.

In this study, we utilized data from a large, multicenter study conducted in multiple LMICs. As such, the results of the study can be considered as generalizable. We also considered non-zero cost for both direct and indirect cost to gain valuable insights considering the overall picture. The costs were adjusted for inflation and converted into I$ for better comparability. However, there were several limitations also. As the GEMS data was collected in 2011-2012, it may fail to capture the current trends. Additionally, cost data was reported by the caregivers which may have caused recall bias. Differences in healthcare systems, currencies, and purchasing power across regions may have complicated the direct comparisons of costs.

## Conclusion

Despite the global decline in pediatric diarrhoeal diseases, they persist as a critical public health issue in LMICs, imposing severe economic strains on households. This study reveals a significant disparity, with indirect costs often surpassing direct costs, underscoring the overlooked financial toll of lost productivity. Regional variations in costs, driven by factors such as household size, maternal education, and access to safe drinking water, highlight the necessity for tailored strategies. Policymakers must prioritize targeted interventions, including bolstering healthcare financing systems, improving access to affordable medications, investing in comprehensive WASH initiatives and promoting timely healthcare-seeking to reduce the economic burden.

## Acknowledgment

We are grateful to the staff, parents, and children of the GEMS for their contributions. The GEMS study was supported, in whole or in part, by the Bill & Melinda Gates Foundation (Grant no. OPP1033572). The conclusions and opinions expressed in this work are those of the author(s) alone and shall not be attributed to the Foundation. Please note works submitted as a preprint have not undergone a peer review process.

The authors of the current study works for icddr,b. The current donors providing unrestricted support to the icddr,b include the governments of Bangladesh and Canada. We thank our core donors for their support and commitment to icddr,b’s research efforts.

## Data availability

Data is available in a public, open access repository. The study used data from The GEMS study which is publicly available at https://clinepidb.org/ce/app/record/dataset/DS_841a9f5259.

## Funding

The authors received no specific funding for the current work

## Conflict of Interest

The authors declare that they have no competing interests.

## Author contributions

Conceptualization: MFAF, TA, SN; Data curation: MFAF, RA; Formal analysis: MFAF, MRI; Interpretation: RA, MFAF, ASGF; Methodology: MFAF, MRI, RA, ASGF; Supervision: ASGF; Writing—original draft: MFAF, MRI, RA; Writing—review and editing: TA, SN, ASGF

---

**S1 Fig.**
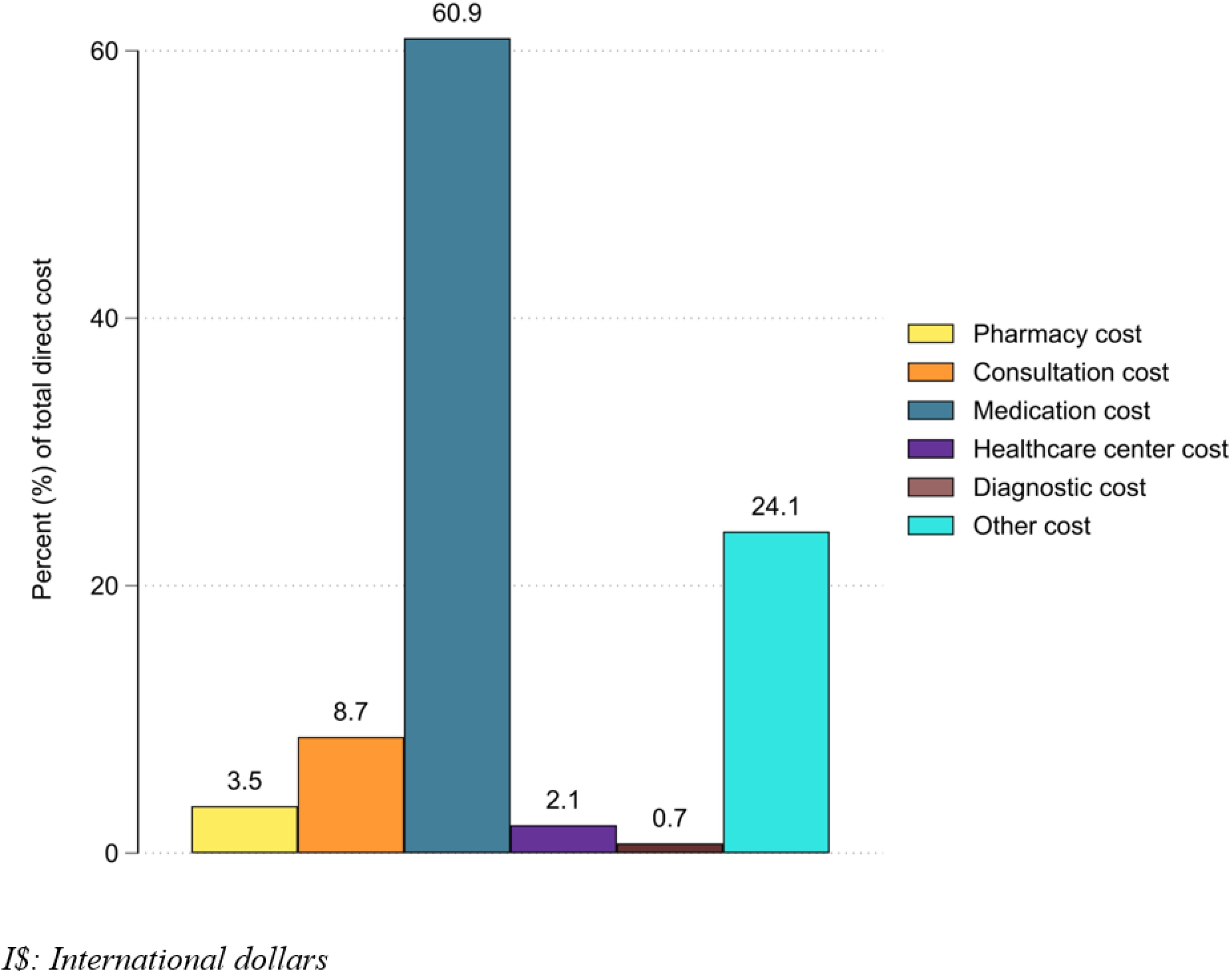
Proportion of total direct medical costs.

**S1 Table.** CPI and PPP of the study sites.

| Country | CPI |  |  | PPP in 2022 |
| --- | --- | --- | --- | --- |
|  | 2011 | 2012 | 2022 |  |
| Bangladesh | 111.4 | 118.3 | 215.9 | 31.37 |
| India | 108.9 | 119.2 | 205.3 | 22.91 |
| Pakistan | 111.9 | 122.8 | 262.6 | 44.10 |
| Kenya | 114.0 | 124.7 | 228.7 | 42.93 |
| Mali | 103.0 | 108.4 | 124.4 | 206.49 |
| Mozambique | 111.2 | 114.1 | 221.4 | 23.55 |
| The Gambia | 104.8 | 109.3 | 219.1 | 17.80 |

495 *CPI: Consumer price index; PPP: Purchasing power parity.*

## Notes

### Competing Interest Statement

The authors have declared no competing interest.

### Funding Statement

This study did not receive any funding

### Author Declarations

The study used publicly available data from: https://clinepidb.org/ce/app/workspace/analyses/DS_2a6ace17a1/new/variables/PCO_0000024/ENVO_00000009

